# Clinical prediction of pathogenic variants in non-coding regions of the human genome

**DOI:** 10.1101/2022.02.25.22271514

**Authors:** Ben O. Petrazzini, Fernando López-Bello, Hugo Naya, Lucia Spangenberg

**Affiliations:** The Charles Bronfman Institute for Personalized Medicine, Icahn School of Medicine at Mount Sinai, New York, NY, USA; Department of Genetics and Genomic Sciences, Icahn School of Medicine at Mount Sinai, New York, NY, USA; Bioinformatics Unit, Institut Pasteur de Montevideo, Montevideo, Uruguay; Pedeciba Bioinformatica, Universidad de la República, Montevideo, Uruguay; Departamento de Producción Animal y Pasturas, Facultad de Agronomía, Universidad de la República, 12900, Montevideo, Uruguay; Departamento de Medicina, Hospital de Clínicas, Universidad de la Republica, Av. Italia, 11600, Montevideo, Uruguay

**Author notes:** **Correspondence:** Lucia Spangenberg, PhD, Mataojo 2020, Bioinformatics Unit, Montevideo, MVD-11400.

## Abstract

Whole genome sequencing has become a wide-spread diagnostic tool for rare diseases patients. This broadens analyses to non-coding regions of the genome showing strong evidence of clinical significance for human Mendelian diseases. Notwithstanding its importance, current *in-silico* prediction tools are restricted to coding sequences which limits its applicability. Additionally, lack of power in discriminating variants of uncertain significance (VUS) limits its clinical utility. Here we present PANCO, a genome-wide pathogenicity prediction tool aiming at reclassification of VUS with a rigorous imputation workflow adapted for non-coding variants. PANCO integrates functional, evolutionary and population frequency information to capture emerging biological signals correctly reclassifying VUS. Importantly, PANCO shows remarkable power in an external validation set, on VUS (AUROC=0.99 and AUROC=0.89, respectively).

## Introduction

The World Health Organization estimates 260M to 400M cases of rare diseases (RD) worldwide^1^, the majority of which affects children^2,3^. On average, patients visit 7 specialists over 8 years awaiting diagnosis, a true “diagnostic odyssey”. The advent of next generation sequencing technologies has facilitated diagnosis over conventional approaches^4^. The current diagnosis rate is approximately 50%; however, this decreases dramatically when a first genetic evaluation fails. Whole genome sequencing (WGS) has the potential to increase the diagnostic yield^5-9^ by incorporating non-coding regions into current pipelines. Untranslated regions (UTRs), introns, regulatory elements and intergenic regions are showing increasing evidence of clinical significance for human mendelian diseases^10,11^ by disrupting splicing^12^, RNA stability^13^, chromatin interactions^11^, transcription^14^ and translation^15^.

*In-silico* prediction methods have facilitated diagnosis in clinical settings; current ACMG guidelines consider such predictions for variant classification^16^. However, the applicability of most methods is restricted to coding regions of the human genome. With the advent of WGS technologies and the ever more evident clinical significance of non-coding regions, there is a need in the field for clinically applicable genome-wide computational tools.

Non-coding regions of the genome have long been beyond the reach of conventional workflows. Scarce functional consequences, together with a lack of statistical power in suspected pathogenic variants and the difficulty of identifying affected protein/s makes it hard to assess their physiological impact. In consequence, the vast majority of non-coding variants are now-a-days classified as variants of uncertain significance (VUS). *In-silico* prediction tools have the potential to overcome this issue by capturing novel signals derived from diverse axes of information. Therefore, clinically useful genome-wide prediction tools are needed to tackle the discrimination of VUS.

Here we present PANCO (PAthogenicity prediction of NOn-Coding variants), a clinically useful *in-silico* tool that accurately discriminates VUS in non-coding regions of the human genome. PANCO presents a rigorous missing value imputation workflow to tackle poorly annotated databases, a longstanding issue when dealing with non-coding regions. PANCO shows remarkable discrimination and case prediction power (area under the receiver operator characteristic curve AUROC=0.99 and Sensitivity=0.99) in an external validation set using variants unseen by its constituent features; bypassing circular overfitting seen in most prediction methods. Additionally, PANCO shows a strong ability to reclassify VUS (AUROC=0.89), underlying it’s potential to aid variant prioritization in clinical settings.

We hope the prioritizing system presented in this paper will help in the interpretation of non-coding variants found in patients with RDs, fostering the slow and harmful diagnosis process^5^.

## Methods

### Data gathering

A total of 490,571 variants with associated clinical significance were obtained from ClinVar^17^ (release March 2019). For each ClinVar variant a combination of clinical significance labels is expected presenting multiple user’s submissions with different experimental evidence. To obtain a well-defined dataset, variants annotated with following labels were considered: Benign, Likely benign, Uncertain significance, Likely pathogenic, Pathogenic and combinations such as Benign/Likely benign, Uncertain significance/Benign, Uncertain significance/Pathogenic and Likely pathogenic/Pathogenic.

### Annotation

ANNOVAR^18^ software was used to annotate all variants with the following databases: refGene (v. 16-4-2018), cytoBand (r. 20-11-2018), ExAC (r. 20-11-2018), avsnp (r. 20-11-2018), dbnsfp (r. 20-11-2018), regsnpintro (r. 8-5-2019), GWAVA (r. 8-5-2019), gnomAD (r. 8-5-2019), Intervar (r. 8-5-2019), Kaviar (r. 8-5-2019), ClinVar (r. 8-5-2019) and Eigen (r. 20-8-2019). This resulted in 107 annotations, most of which use functional information and are therefore not relevant for non-coding variants. Features restricted to coding variants were removed, as well as features with high correlation (Pearson’s > 0.8), resulting in a reduced set of 11 annotations to include in the ML workflow (DANN^19^, fathmm.MKL^20^, fitCons^21^, GERP^22^, phyloP20^23^, phastCons20^24^, regSNP^25^, GWAVA^26^, gnomAD^27^, Kaviar^28^ and Eigen^29^).

### Missing values imputation

Missing annotations are a longstanding issue when working with non-coding variants. Missing values are usually dealt with by taking the feature’s average/median. However, this carries no biological significance, as by doing so we tend to underestimate relevant information. For example, a highly pathogenic variable is expected at low population frequencies; however, imputing the average would result in higher frequencies than expected, implying most likely an underestimation of its pathogenicity and further reducing the predictive power of our method. To tackle this issue, several predictive imputation methods were tested in our dataset. These are a kNN-based method from *DMwR*^30^ (v. 0.4.1) package, a flexible additive model from *Hmisc*^*31*^ (v. 4.2-0) package, a function working over chained equations of Markov models from *mice*^32^ (v. 3.6.0) package, a regression model from *mi*^33^ (v0.10-2) package and a maximum-likelihood method from *Amelia*^34^ (v. 1.7.5) package, using the NAsImpute^35^ package available in R.

To evaluate the performance of each imputation method 600 fully annotated data points in each column were randomly masked (turn to missing value), then imputed them using each method and compared the result to the originally observed values. This process was iterated 100 times on each of the 11 columns in our dataset. Root Mean Squared Error (RMSE) and Mean Absolute Error (MAE) metrics were used to determine error metrics between imputed and observed values for each method over each column. The reported values were scaled by the columns range for comparison purposes.

None of the evaluated methods produced an accurate enough prediction for the regSNP column. Given that the latter is a useful pathogenicity predictor for non-coding mutations, a Neural Network-based approach was developed to predict its unobserved data. regSNP was taken as the model’s output vector; a Multilayer Perceptron was then fitted to the remaining 10 fully annotated features in our training set. The resulting model was then used to predict missing values in regSNP. To account for biases, the output vector (regSNP) was balanced by reducing the number of variants with a pathogenicity score between 0 and 0.1 (Supplementary Figure 1).

To ensure our annotations are derived from biological information and not imputed values, only variants with less than 66% missing values were considered.

### Machine learning workflow

A number of machine learning (ML) algorithms were tested; only the best performing algorithm, Gradient Boosted Trees^36^, was used to fit the final model.

The ML workflow is summarized in Figure 1. To account for the model’s bias, 4,000 Pathogenic and 4,000 Benign variants were used for training. The rest were used for testing, which included 421 Pathogenic, 4,075 Pathogenic/Likely Pathogenic, 3,170 Uncertain Significance, 4,415 Benign/Likely Benign and 1,092 Benign variants left for testing. To generate a balanced test set, 421 Pathogenic variants and an equal number of Benign variants were selected. The selection was repeated 100 times to avoid sampling biases. Reported performance metrics in the test set correspond to the mean and standard deviation across those 100 iterations.

**Figure 1.**
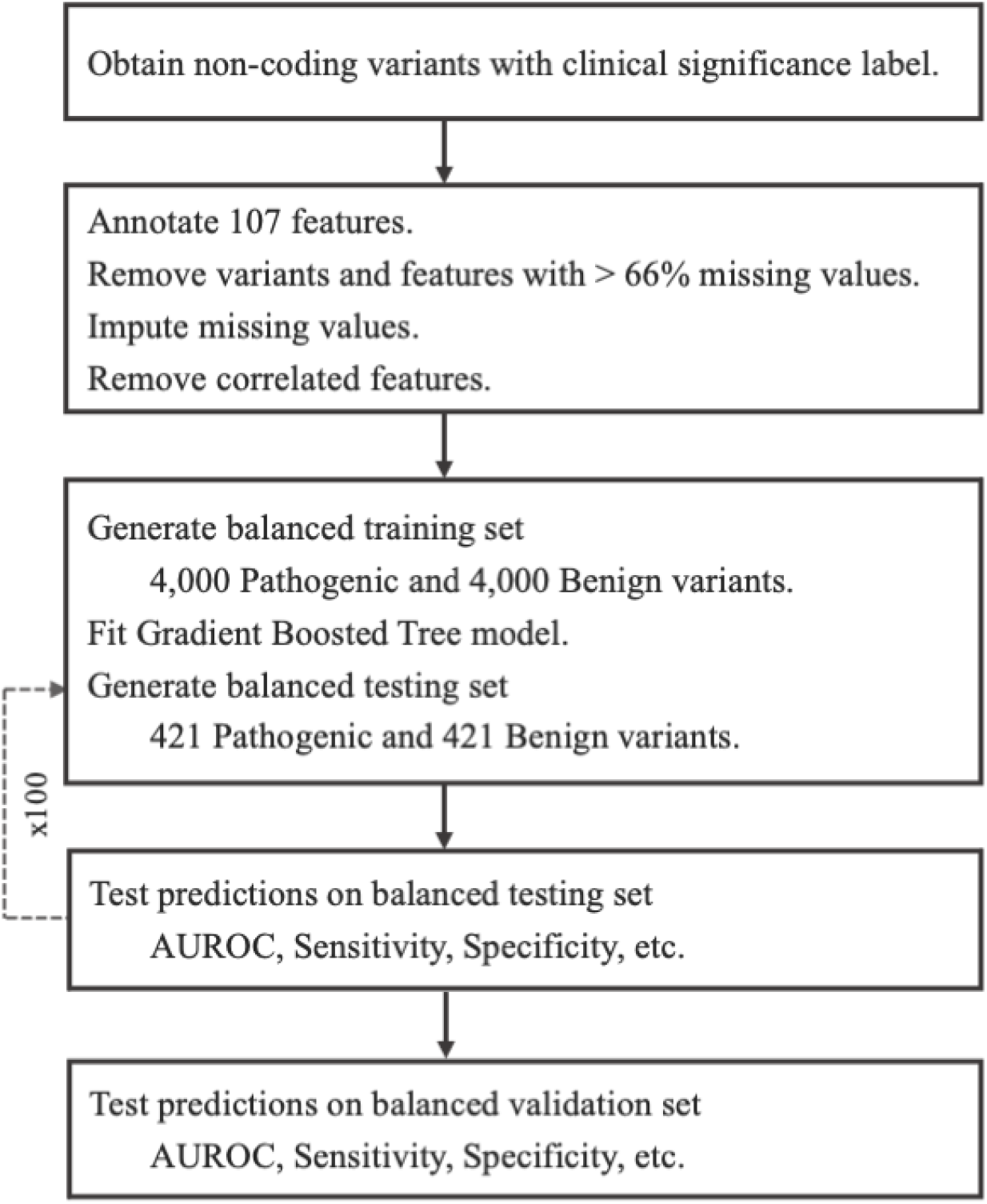
Workflow AUROC corresponds to area under the receiver operator characteristic curve.

A 10-fold cross-validation approach was performed within the training set to determine the best-performing parameters. Performance metrics were subsequently computed in the training set to evaluate overfitting when comparing with the test set. Overall accuracy, sensitivity, specificity and area under the receiver operator characteristic curve (AUROC) were used. The *caret*^37^ (v. 6.0-84) package and roc from *pROC*^38^ (v. 1.15.3) package were used to fit the models and compute performance metrics. The fitted model was then used to compute performance metrics on the above-mentioned unseen test data. Two approaches were taken to determine if PANCO was able to reclassify VUS. First, 3,170 VUS were predicted to evaluate how PANCO is able to cluster these variants with a high degree of confidence. Second, a secondary test set was created by taking variants that were labeled as VUS in the first place and then reclassified to either Benign or Pathogenic in ClinVar. This secondary test set is then composed of variants carrying the same characteristics of VUS that are hard to classify, but with a posterior curated clinical significance label allowing us to compute performance metrics.

This way of evaluating our two models in three different steps has proven to be a good approach to assess their performance in different kinds of data, going from variants with clearly defined pathogenicity, to somewhat defined variants and non-labeled variants.

### External validation

Variants from additional sources were used to generate an external validation set. A big issue when evaluating variant classification models is that the scores used for training are themselves trained on the variants used for testing. This circular overfitting makes it hard to obtain a true external validation set to measure true performance metrics. To overcome this issue, two sets of variants novel to PANCO’s constituent features at the moment the model was fitted were used as external validation; these are the latest version of ClinVar (release June 2020) and a recently released dataset from a Japanese population (GEM)^39^. The latest release of ClinVar was used to obtain Pathogenic non-coding variants with evidence of pathogenicity, which are hard to find. A random sample of non-coding variants with allele frequency >1% in the GEM dataset and not present in gnomAD were included in the validation set as Benign variants. These were then predicted using PANCO to compute performance metrics on an external validation set. To generate a balanced validation set, all available Pathogenic variants and an equal number of Benign variants were selected. This was repeated 100 times to avoid sampling biases. Reported performance metrics in the external validation set correspond to the mean and standard deviation across those 100 iterations.

## Results

### Data gathering

97,736 non-coding variants with classic clinical significance labels (Pathogenic, Likely pathogenic, Uncertain Significance, Benign and Likely Benign) were downloaded from ClinVar. Of those, 21,173 variants had under 66% missing values (features) with following clinical significance labels: 4,421 Pathogenic, 4,075 Pathogenic/Likely Pathogenic, 3,170 Uncertain Significance, 4,415 Benign/Likely Benign and 5,092 Benign.

### Imputation

As expected, all imputation methods outperformed the imputation-by-average approach in every column (Supplementary Table 1). Interestingly, the k-NN-based method had the best performance in 8 out of the 11 imputed features, these are DANN, fathmm, fitCons, GERP, phastCons20, GWAVA, gnomAD and Eigen as previously reported^35^. For the remaining features, the regression model was the best at imputing phyloP20 and Kaviar. As detailed above, a Neural Network approach was developed to deal with missing values in regSNP.

Given these results, *knnImputation* was used to predict the 8 above-mentioned features, whereas *mi* was used to predict phyloP20 and Kaviar and our Neural Network-based approach was used to predict regSNP missing values.

### PANCO identifies signals of pathogenicity in non-coding variants

The best performing algorithm was Gradient Boosted Trees from *xgboost* package^36^ (v. 0.90.0.2). The parameters used to fit this model were *nrounds = 400, max_depth = 5, eta = 0*.*2, gamma = 0, colsample_bytree = 0*.*4, min_child_weight = 1* and *subsample = 0*.*8*.

PANCO shows strong power in discriminating both Pathogenic and Likely Pathogenic variants from Benign and Likely Benign variants in internal blind test sets, as well as an external validation set. When predicting a blind set of Pathogenic and Benign variants, PANCO shows similar performance to the training set (AUROC=0.99 and 0.99 for the test and train set, respectively) (Figure 1A, Figure 1B and Table 1) indicating minimal overfitting. When predicting a blind test set of Likely Pathogenic and Likely Benign variants, PANCO shows similar discrimination power (AUROC=0.99) with a small decrease in case prediction power (Sensitivity=0.96) (Figure 1C and Table 1). This suggests PANCO captures pathogenicity signals in presumably weaker variants, although in a smaller proportion compared to Pathogenic variants. Finally, PANCO shows a performance in the external validation set comparable to its performance in the internal blind test set (AUROC=0.99 for both the external validation and internal blind test set) (Figure 1A and Figure 1D), confirming the minimal overfitting seen above, and suggesting PANCO will perform similarly in novel external dataset provided by any user.

**Table 1.** Performance metrics

| **Dataset** | **AUROC** | **Accuracy** | **Sensitivity** | **Specificity** | **PPV** | **NPV** |
| --- | --- | --- | --- | --- | --- | --- |
| Train | 0.99 | 0.99 | 0.99 | 0.99 | 0.99 | 0.99 |
| Test | 0.99 (0.00) | 0.99 (0.01) | 0.99 (0.01) | 0.99 (0.00) | 0.99 (0.00) | 0.99 (0.01) |
| Test on LB/LP | 0.99 (0.00) | 0.97 (0.00) | 0.96 (0.00) | 0.99 (0.00) | 0.99 (0.00) | 0.96 (0.00) |
| Validation | 0.99 (0.00) | 0.99 (0.00) | 0.99 (0.00) | 0.99 (0.00) | 0.99 (0.00) | 0.99 (0.00) |

### PANCO identifies novel signals in variants of Uncertain Significance in non-coding regions

Classifying VUS have shown to be a daunting task for current prediction tools. A combination of mixed clinical evidence and ambiguous characteristics hinders ascertainment of these variants. Two separate analyses were undertaken to evaluate PANCO’s performance in this task.

First, we examined whether PANCO is able to cluster (into categories, such as Benign/Pathogenic) a large number of VUS with high confidence. A preliminary PCA analysis on 11 features shows that VUS cluster together within two groups: benign and pathogenic variants. (Figure 3A). This indicates that vectorial transformations in our dataset are able to unravel novel signals for VUS; information that is non-detectable when analyzing individual annotations and is common to either Pathogenic or Benign variants. The interaction between features driving this signal was captured by a Gradient Boosted Tree algorithm, suggesting a potential to cluster/classify VUS. This is further confirmed when looking at the distribution of prediction scores for all VUS (Figure 3B). Two distinct clusters can be observed, each one with a preponderant number of high-confidence annotations, meaning PANCO is over 95% confident that the variant is either Pathogenic or Benign. These results suggest PANCO is capturing novel information that allows for a distinct clustering of VUS, presumably obtaining information used to classify Pathogenic or Benign variants.

**Figure 2.**
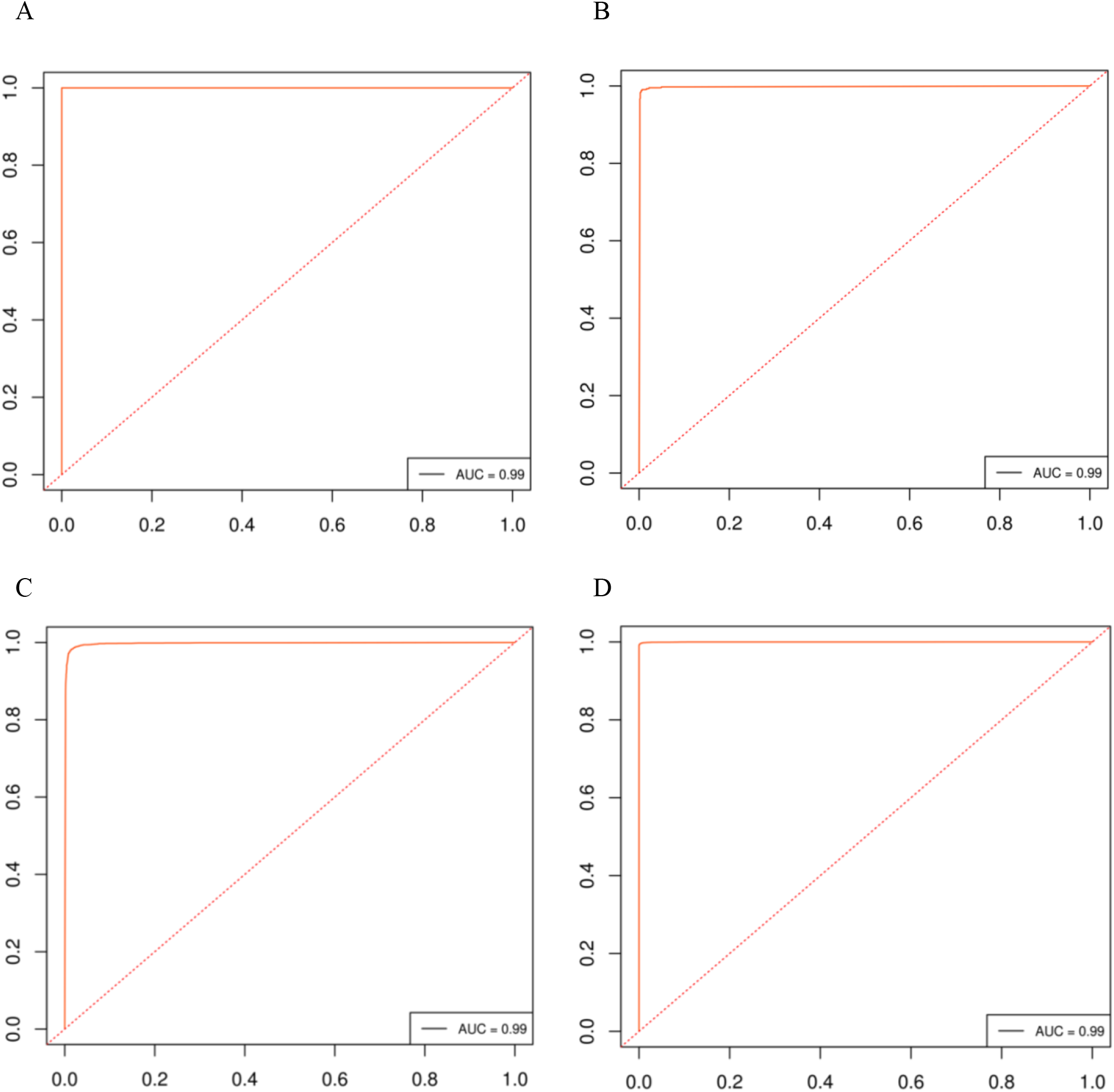
Receiver operator characteristic curves A) Receiver operator characteristic curve on the train set. B) Receiver operator characteristic curve on the test set. C) Receiver operator characteristic curve when testing Likely Pathogenic and Likely Benign variants. D) Receiver operator characteristic curve on the external validation set. Y and X axis correspond to averaged true positive and false negative rates respectively across 100 iterations. AUC corresponds to the area under the curve.

**Figure 3.**
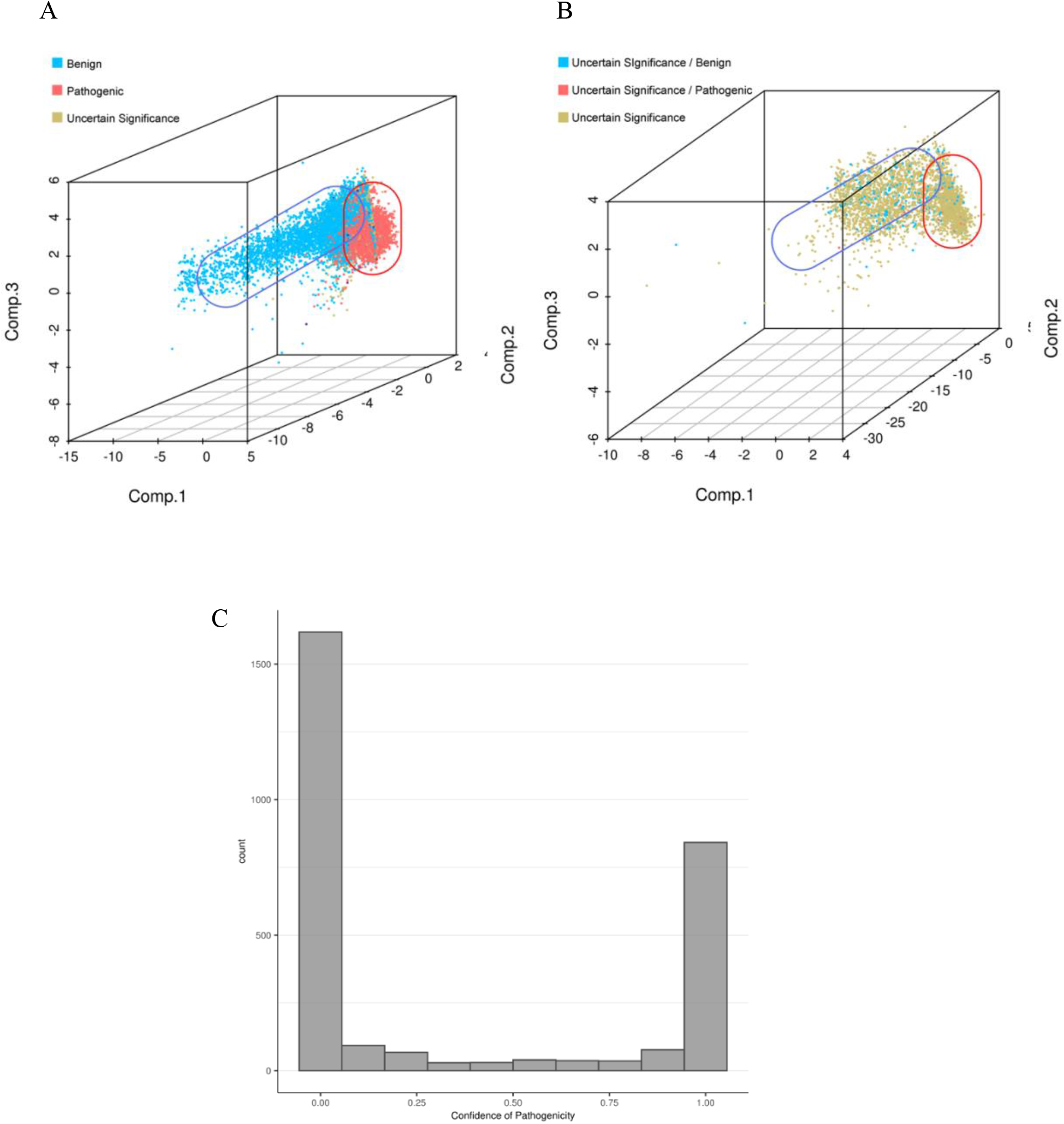
Clustering of Uncertain Significant variants. A) Principal component analysis on all variants (left) and Uncertain Significance, Uncertain Significance / Benign and Uncertain Significance / Pathogenic variants (right). B) Distribution of predicted PANCO scores for all Uncertain Significance variants.

Second, we assessed PANCO’s performance on variants originally classified as VUS in ClinVar and later ascertained towards Benign or Pathogenic. These are presumed to have similar characteristics to current VUS but have currently a clinical significance label (Pathogenic/Benign), allowing to compute performance metrics. PANCO shows remarkable performance at discriminating these variants (AUROC=0.89) (Figure 4A). On a case/control analysis it shows decreased performance for the positive class, indicating it underestimates pathogenic load for 37% of Pathogenic variants previously labelled with VUS (Figure 4C). However, the strong discrimination performance and high-confidence at calling Benign variants previously labelled VUS (NPV=0.85) indicates more Pathogenic variants can be identified by doing a sensitivity analysis.

**Figure 4.**
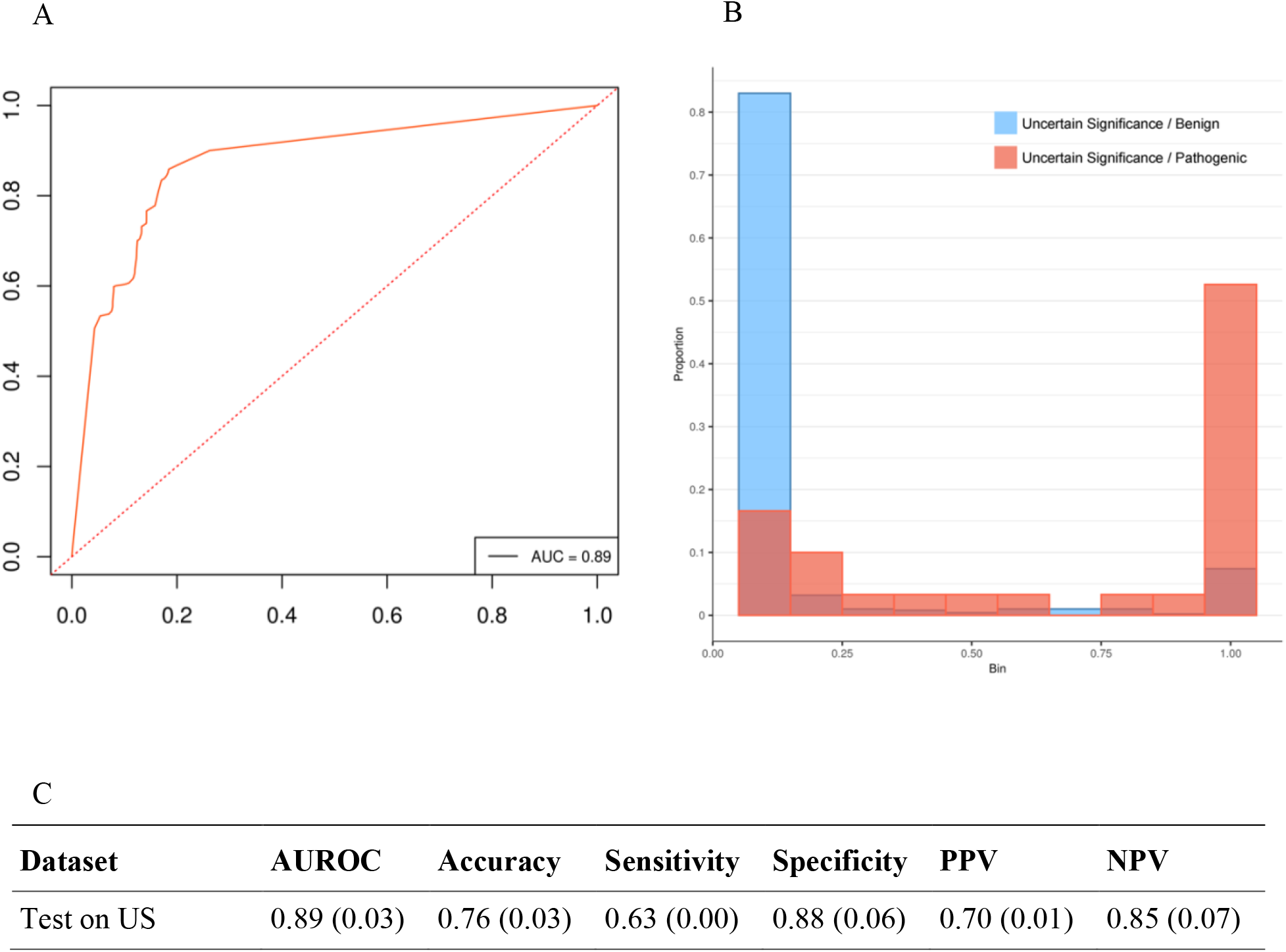
Distribution of PANCO’s score predicting Uncertain Significant variants. A) Receiver operator characteristic curve when testing Uncertain Significance variants. Y and X axis correspond to averaged true positive and false negative rates respectively across 100 iterations. AUC corresponds to area under the curve. B) Distribution of predicted Uncertain Significance / Benign and Uncertain Significance / Pathogenic variants. Y axis corresponds to the proportion of variants classified by PANCO in each of the bins shown in the X axis. C) Performance metrics when testing Uncertain Significance variants. AUROC corresponds to area under the receiver operator characteristic curve, US corresponds to Uncertain Significance variants.

Overall, results from both analyses suggest PANCO can identify novel signals in non-coding VUS that could help reclassifying them as either Pathogenic or Benign.

### Three features account for the majority of the model’s weight

A feature importance analysis reveals that regSNP, FATHMM and DANN are driving the prediction (Figure 5). These three features account for 80.4% of the model’s weight, and are the only ones carrying functional information. regSNP is a random forest-based model integrating functional (RNA splicing and protein structure) and evolutionary features, which accounts for 37.4% of the model’s weight. FATHMM-MKL and DANN are both machine learning approaches using multiple kernel learning and deep neural networks, respectively. Both include functional information such as histone modification, open chromatin, transcription factor binding sites and others, and account for 21.6% and 21.3% of the model’s weight, respectively.

**Figure 5.**
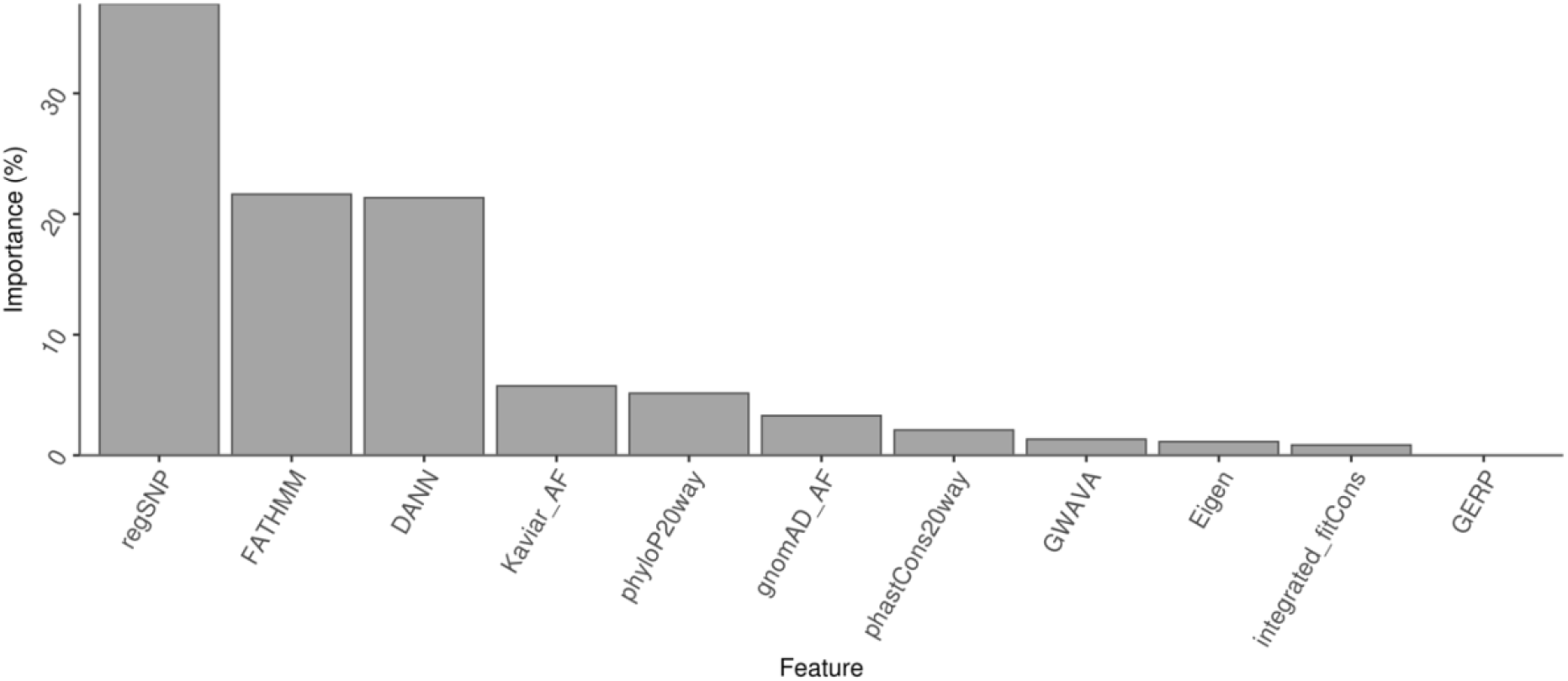
Feature importance The feature importance corresponds to the percentage of contribution from that feature to fitting the final model.

## Discussion

Given the large number of missing annotations in non-coding regions, a rigorous imputation workflow was performed in this study. Several imputation algorithms were tested on our dataset using the NAsImpute method. Different observed data is better fitted to different algorithmic frameworks^35^. Biological information is better preserved when applying a rigorous imputation workflow, ensuring robust annotations with respect to the model’s outcome which would ultimately result in a boost of power when fitting any given ML model.

PANCO shows a significant improvement in prediction power compared to current methods targeting non-coding variants. This performance can be generalized to any new variant, as it was computed on a novel set of Benign and Pathogenic variants not seen by any of PANCO’s constituent features. A trustworthy report of performance metrics is extremely important to implement prediction models in clinical practice. Most current methods, however, report their performance metrics on variants not present in their training or testing set but seen by the model’s constituent features; this results in overestimated performance metrics. A tool reporting overestimated performance metrics will, in consequence, provide misleading information for clinicians struggling with rare disease diagnosis. It is therefore extremely important to rigorously validate the model using novel variants that diverge from this circular overfitting.

Efforts have been made over recent years to develop tools for the interpretation of VUS in coding regions^40,41^. However, the vast majority of non-coding variants in a rare disease patient will not meet requirements from the ACMG for a Benign or Pathogenic classification. Therefore, the primary clinical utility of prediction tools tackling non-coding regions is to accurately discriminate VUS with high confidence. An analysis using manually curated Uncertain Significance / Benign and Uncertain Significance / Pathogenic variants shows PANCO could aid ascertainment in these variants with strong discrimination power.

Pathogenicity signal is likely to be a composite of evolutionary, functional and frequency information. While evolutionary and frequency information can be mapped throughout the genome, only a reduced set of functional features can be used to characterize non-coding variants. Therefore, prediction tools integrating substantial functional information for non-coding regions are likely to capture additional axes of pathogenicity signals. Consequently, PANCO’s prediction heavily accounts for RNA splicing, CG content, open chromatin, histone modifications, footprints and binding sites. Importantly, PANCO also integrates protein structure that is contained in regSNP by mapping to the closest exome, an approach that has recently been validated^42^. regSNP being the most important feature not only shows that functional information improves pathogenicity prediction in non-coding variants but also validates our neural network imputation approach (see Methods); underlying the importance of appropriately imputing missing values.

In conclusion, PANCO can accurately discriminate pathogenic variants by exploiting functional, evolutionary and frequency information from non-coding regions of the human genome. It’s unique set of features strengthens scarce functional information for non-coding variants, allowing for precise discrimination of variants of uncertain significance. This carries strong clinical utility, which is why PANCO can serve as a variant prioritization tool aiding clinical geneticists to identify functional mutations in ambiguous regions of the genome.

Some of the limitations of our study includes: i) it is restricted to 21,173 variants with pathogenicity label from ClinVar. ii) PANCO’s performance in VUS is derived from a low number of Uncertain Significance / Pathogenic variants. These two limitations are part of a bigger burden in the field of human genetics; a shortage in resources with clinical interpretation of non-coding variants. iii) Calculating PANCO’s score could be computationally intensive, which could limit its clinical applicability in hospital settings with limited resources. To aid the implementation of this tool we provide precomputed pathogenicity estimations for 21M non-coding variants from the dbSNP, available at https://github.com/OmegaPetrazzini/PANCO.

## Supporting information

Supplemental methods and results.

## Data Availability

All data produced are available online at

https://github.com/OmegaPetrazzini/PANCO

## Author Contributions

Dr. Spangenberg and Mr. Petrazzini had full access to all of the data in the study and take responsibility for the integrity of the data and accuracy of the data analysis.

*Concept and design:* Petrazzini and Spangenberg.

*Acquisition, analysis, or interpretation of the data:* All authors.

*Drafting of the manuscript:* Petrazzini and Spangenberg.

*Critical revision of the manuscript for important intellectual concept:* All authors.

*Statistical analysis:* Petrazzini and Spangenberg.

*Administrative, technical, or material support:* Petrazzini, Naya and Spangenberg.

*Supervision:* Spangenberg.

## Disclaimer

The content is solely the responsibility of the authors.

## Data sharing statement

Not applicable.

