## Supplemental methods and results. for "Clinical prediction of pathogenic variants in non-coding regions of the human genome"

Figures

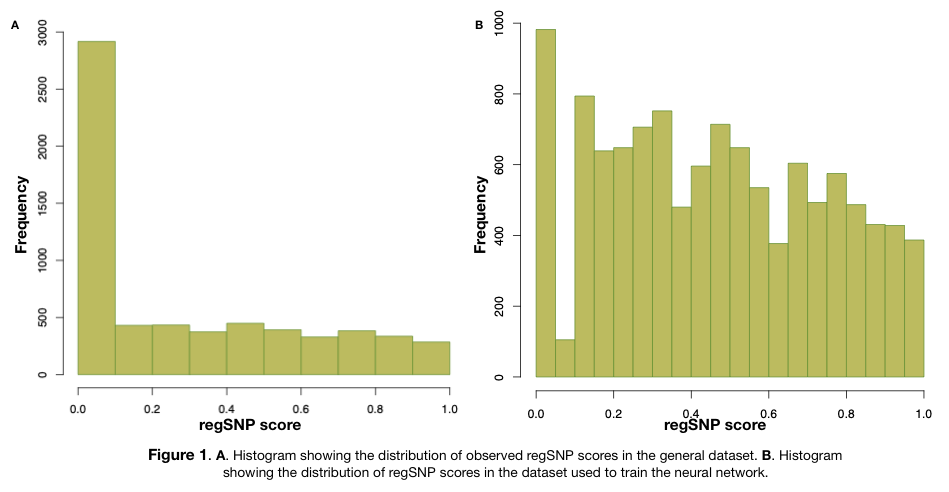
Supplementary Figure 1. Distribution of regSNP scores.

A) Histogram showing the distribution of regSNP in the entire dataset. B) Histogram showing the distribution of regSNP used to fit the neural network used for imputation.

Tables

Supplementary Table 1.

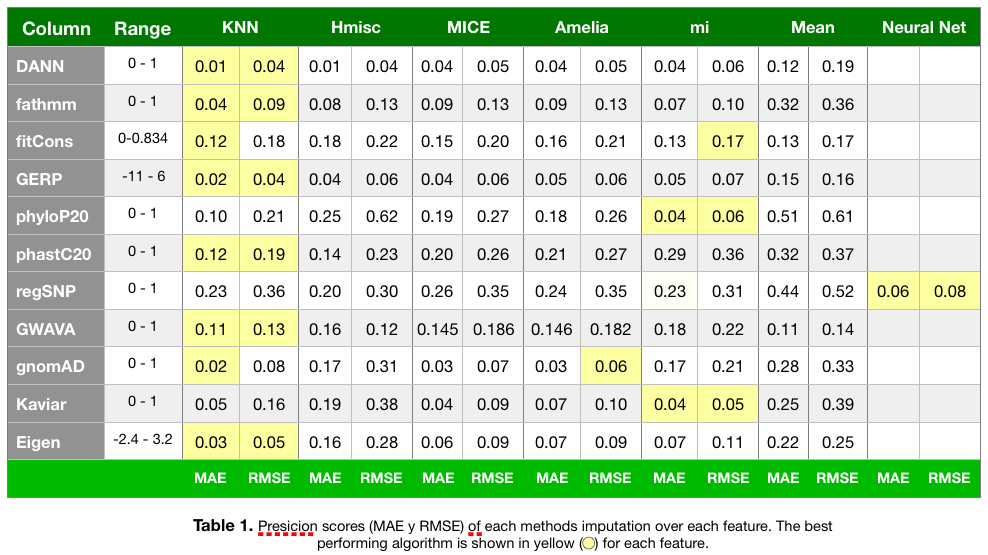
